# Effect of Daytime Nap on Attention, Working Memory, and Recognition Memory

**DOI:** 10.1101/2023.08.14.23294066

**Authors:** Shahab Zare, Fariborz Dortaj, Javad Setareh

## Abstract

Daytime napping has gained attention for its potential to enhance cognitive functions. While the positive impact of nocturnal sleep on cognitive functions is widely acknowledged, limited research has focused on the effects of daytime naps. In this study, we investigated the influence of a 30-minute afternoon nap on cognitive functions in healthy adults.

Ten participants (4 females) were involved in both nap and non-nap (control) conditions. Participants were randomly selected from the sleep department of Zare Hospital. During the nap condition, participants engaged in a 30-minute nap at approximately 14:00. Cognitive assessments were performed 30 minutes after the nap to reduce the effects of sleep inertia. Three days after nap condition, cognitive evaluations were conducted again at approximately 15:00, in the non-nap condition.

Working memory, evaluated using the Backward Digit Span Task using a paired sample t-test, exhibited significant improvement during napping (p = 0.0164, d = 0.459), highlighting its practical significance. Attention, assessed with the Stroop test, and the Wilcoxon signed-rank test showed significant differences between nap and non-nap conditions, emphasizing napping’s role in sustaining attention and alertness. Recognition memory, gauged through the Auditory Verbal Learning Test, revealed superior performance in the nap condition, particularly in False Alarm scores.

Despite limitations such as sample size and nap monitoring, our study contributes valuable insights into the cognitive benefits of daytime napping. By refining our understanding of napping’s effects, tailored interventions can be developed, promoting cognitive health and performance enhancement across various contexts.

Iranian Registry of Clinical Trials (IRCT) registration number 66862. Date of registration: 12/31/2022

The study was approved by the Research Ethics Committee of Allameh Tabataba’i University (IR.ATU.REC.1401.030).

**Statements and Declarations:** The writers confirm that they have no affiliations or engagements with any organization or institution that holds any financial or non-financial interest in the subject matter or materials addressed in this publication.

## 1. Introduction

Circadian rhythms are approximately 24-hour physiological patterns that harmonize internal states with the external environment and anticipate various physiological needs that emerge throughout the day. During the mid-afternoon siesta or nap period, a time when many individuals opt for a nap, the circadian signal promoting wakefulness experiences a noticeable decline. This circadian wakefulness signal fades as bedtime approaches, ultimately leading to the onset of sleep (1). Nine to 15% of adults with chronic sleep disturbances report daytime effects of their poor sleep quality. In a large-scale study conducted in the USA, 34.8% of adults reported having insufficient sleep (2). Sleep deprivation can result in fatigue, daytime drowsiness, and impaired memory function. Sleep deprivation has been linked to a variety of mental and physical health issues, including anxiety, depression, and cardiovascular disease (3).

Considering the widespread occurrence of sleep deprivation in our rapidly evolving modern society, scheduled naps are gaining traction as a potential remedy for countering the adverse cognitive consequences associated with chronic, fragmented nighttime sleep (4). There is plenty of evidence to suggest that sleep is essential for maintaining cognitive function (5). The advantages of a nap may vary according to the degree of sleep deprivation experienced beforehand. Research has indicated that engaging in an afternoon nap during a period when individuals are prone to drowsiness can alleviate homeostatic sleep pressure and enhance levels of alertness and memory (6, 7, 8, 9, 10). Adults and adolescents both benefit from afternoon naps in terms of memory (9, 11, 12, 13, 14).

In infants, naps closely resemble nighttime sleep due to their abundance of rapid eye movement (REM) sleep (15). As children enter early childhood, naps primarily consist of non-rapid eye movement (NREM) sleep with minimal REM sleep (16). During young adulthood, longer naps can encompass both NREM and REM sleep cycles (17). In the case of older adults, naps are characterized by a prevalence of lighter NREM sleep stages, a brief period of slow-wave sleep (SWS), and occasionally REM sleep occurring less frequently (18). In the laboratory, taking a nap after learning helps with memory consolidation, whereas taking a nap before learning helps with encoding new information (19, 20, 21, 22).

Attention is crucial in everyday life. The word “attention” has many meanings but typically refers to selectivity in processing. Selective attention (focused attention) is studied by presenting individuals with two or more stimuli at the same time and instructing them to respond to only one. Research on focused or selective attention tells us how effectively we can select certain inputs and avoid being distracted by non-task stimuli (23).

Working memory is a cognitive mechanism that enables us to actively retain small chunks of information for use in ongoing tasks (24). Short-term memory is concerned mainly with storing information for a brief period of time (for example, remembering a phone number), whereas working memory is concerned not just with how information is stored but with how information is manipulated in the service of various forms of cognition (25). Recognition memory is composed of two separate systems, which may be the reason for its overall advantage over recall tests. One process, familiarity, is thought to be independent of context, and the recognized item just has a familiar feeling. The other mechanism, remembrance, depends on the context and entails recalling particular details from the study incident. Tests of recall are thought to be virtually entirely dependent on memory (26). There is plenty of evidence to suggest that sleep is essential for maintaining cognitive function. Deficits in executive function and working memory are some examples of the consequences of sleep deprivation (5).

While poor sleep is associated with a variety of sociodemographic differences, a key risk factor for poor sleep involves individuals’ personality traits—individual differences in persistent patterns of thinking, feeling, and acting (27). Evidence suggests that sleep disturbances are fairly stable over time, which also implies that stable features of individuals, such as personality traits, could be important concomitant factors (28).

Despite the fact that the impact of afternoon naps on cognitive processes has previously been studied, this study attempted to control the influence of personality characteristics by using the same set of participants in both conditions to investigate the effect of a daytime nap on attention, working memory, and recognition memory.

## 2. Methods and Materials

In accordance with best practices for research transparency and in compliance with ethical guidelines, the present study was prospectively registered with the Iranian Registry of Clinical Trials (IRCT) under registration number 66862. This registration was completed prior to the commencement of data collection and serves as a testament to the study’s commitment to rigorous research methodology and reporting.

The study was approved by the Research Ethics Committee of Allameh Tabataba’i University (IR.ATU.REC.1401.030). We confirm that all methods were performed in accordance with the relevant guidelines and regulations.

### 2.1. Participants

A total of 15 healthy participants were recruited for the present study. Participants were randomly selected from the sleep department of Zare Hospital. Participants were eligible if they were between 25 and 50 years old, had no known neurocognitive or sleep disorders, were not shift workers, and had a body mass index between 18.5 and 24.5. Exclusion criteria included the intake of sleeping pills, being colorblind, and having a history of neurological and psychiatric diseases.

### 2.2. Study protocol

In this study, to minimize between-group variation, the participants who took part in the intervention phase (Nap) were also included in the non-intervention phase (Non-nap). Participants were asked to maintain a normal sleep duration (i.e., 7–9 h) throughout the experimental period to avoid any undesirable effects of sleep deprivation. All participants in the study were given a chance to take a 30-minute afternoon nap. They got into bed at 13:45 in rooms that were favorable to sleep (i.e., dimly lit and quiet). After 15 minutes of becoming accustomed to their sleep environment, participants were asked to take a nap. To avoid sleep inertia, 30 minutes after waking up, participants were subjected to cognitive assessments (29, 30). After three days, they participated in the second phase of the study and were cognitively evaluated at 15:00 without experiencing sleep in the afternoon.

### 2.3. Measurements

#### 2.3.1. Working Memory Assessment: Backward Digit Span

The Backward Digit Span Task is a component of the Wechsler Memory Scale that is used for different age groups. Although this subtest primarily assesses auditory working memory, attention can also be evaluated within its framework. In this test, the participant is required to not only hold the numbers in their memory but also to repeat them in reverse order, from last to first, as compared to what was presented to them. The numbers presented range from 1 to 10, and it is worth noting that there is no logical relationship between the numbers in each sequence. If the participant correctly recited the reversed sequence of all the numbers presented in each trial, they receive a score of one. However, if they make a mistake in reciting the numbers, they received a score of zero. After two mistakes in a row, the test is completed, and the sum of the scores (zero or one) is the individual’s final score. This score, along with the length of the longest sequence they were able to correctly repeat, is recorded.

#### 2.3.2. Recognition Memory Assessment: Auditory Verbal Learning Test (AVLT)

Recognition memory assessments typically consist of displaying a diverse array of items to participants, who are then tasked with determining the prior presentation status of each item (frequently characterized by a balanced distribution of 50% previously presented items and 50% novel items) (23). The AVLT evaluates individuals’ abilities to decipher, retain, store, and retrieve verbal information. Two lists of unrelated words were read aloud to participants (the first list was repeated four times and the second list once), and they were instructed to recall as many words as possible immediately after the presentation (immediate recall). After a 20-minute interval, participants were again asked to recall the words (delayed recall), and their performance was assessed by recording the number of correctly recalled words. Then, a list of words, which is a combination of words from the first and second lists and new words, was read to the participants, and they were asked to determine whether the words were in the first list or not (recognition phase). We used the Persian version of the AVLT in this study (31).

#### 2.3.3. Attention Assessment: Stroop Test

The Stroop test was utilized to assess participants’ attention (32). Neutral, congruent, and incongruent stimuli are the three basic categories of stimuli included in the Stroop test. Neutral stimuli (Control trial) include ones that could be composed of colorful forms or color-words written in black ink. Color words printed in the same ink color as the word (for example, “blue” written in blue ink) are considered congruent stimuli, whereas incongruent stimuli are those that have the color-word printed in a different ink color (for example, “blue” written in red ink) (33). The total number of error color identifications and response times were recorded for each participant in each of the three trials. The Stroop test has been widely used in research to assess attentional control and cognitive flexibility, and it has demonstrated good reliability and validity. Therefore, in this study, the Stroop test was selected as a reliable and valid tool for assessing attentional processes.

### 2.4. Statistical analysis

R version 4.3.1 was utilized for statistical analysis (34). Measurement data are presented as the mean and standard deviation, and population data are expressed as the number of cases and/or rate (%). Statistical methods included Shapiro—Wilk, Paired sample t-test, and Wilcoxon signed rank test. Specific approaches are listed in the “Results” section. P <0.05 was considered statistically significant.

## 3. Results

Five participants failed to return for the non-intervention session. Data for these subjects were removed from all analyses. The final sample consisted of 10 participants in total, with 4 females; the overall mean age was 32.1 years (Table 1).

**Table 1.** Demographics of the sample.

|  | Mean | Max | Min | Standard Deviation |
| --- | --- | --- | --- | --- |
| Age (yrs) | 32.1 | 41 | 25 | 5.46 |
| Body Mass Index (kg/m<sup>2</sup>) | 22.28 | 24.4 | 19.8 | 1.46 |
| Education (yrs) | 15.6 | 18 | 12 | 1.83 |

### 3.1. Analysis of Normality

Before conducting further statistical analyses, we performed a test for normality on the data using the Shapiro—Wilk test. For both the non-intervention and with-intervention datasets, we evaluated whether the data significantly deviated from a normal distribution.

The Shapiro—Wilk test was conducted using R, and the function ‘shapiro.test()’ was employed to compute the test statistics and corresponding p-values. The significance level (alpha) was set to 0.05, as is standard in statistical analysis.

The results of the Shapiro—Wilk test indicated that data related to working memory assessment demonstrated p-values greater than the significance level (alpha = 0.05). As a result, we fail to reject the null hypothesis, suggesting that there is no significant evidence to reject the assumption of normality. Otherwise, the results of the normality check indicated that the distribution of attention and recognition memory exhibited significant deviations from a normal distribution. We used the Wilcoxon signed-rank test for data that deviated from a normal distribution, and to calculate the effect size by considering the sample size, we used Hedges’ g.

### 3.2. Working Memory

The paired sample t-test was employed to compare the mean Backward Digit Span scores between the two condition s (Nap and Non-nap). This parametric test revealed a significant difference in Backward Digit Span performance (t = 2.940, p = 0.0164, Hedges’ g = 0.459). Participants demonstrated a higher mean Backward Digit Span score in the situation with a daytime nap (M = 14.30, SD = 2.40) compared to the situation without intervention (M = 12.9, SD = 3.21) (Table 2) (figure 1).

**Table 2.**
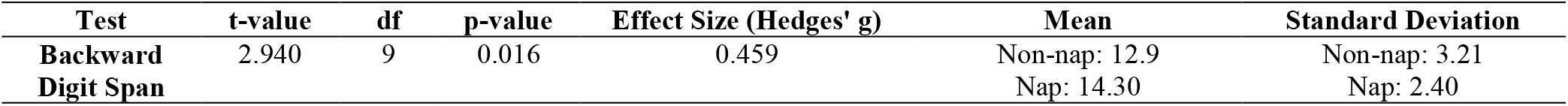
Results of working memory assessment.

| Test | t-value | df | p-value | Effect Size (Hedges' g) | Mean | Standard Deviation |
| --- | --- | --- | --- | --- | --- | --- |
| **Backward Digit Span** | 2.940 | 9 | 0.016 | 0.459 | Non-nap: 12.9 | Non-nap: 3.21 |
|  |  |  |  |  | Nap: 14.30 | Nap: 2.40 |

**Figure 1.**
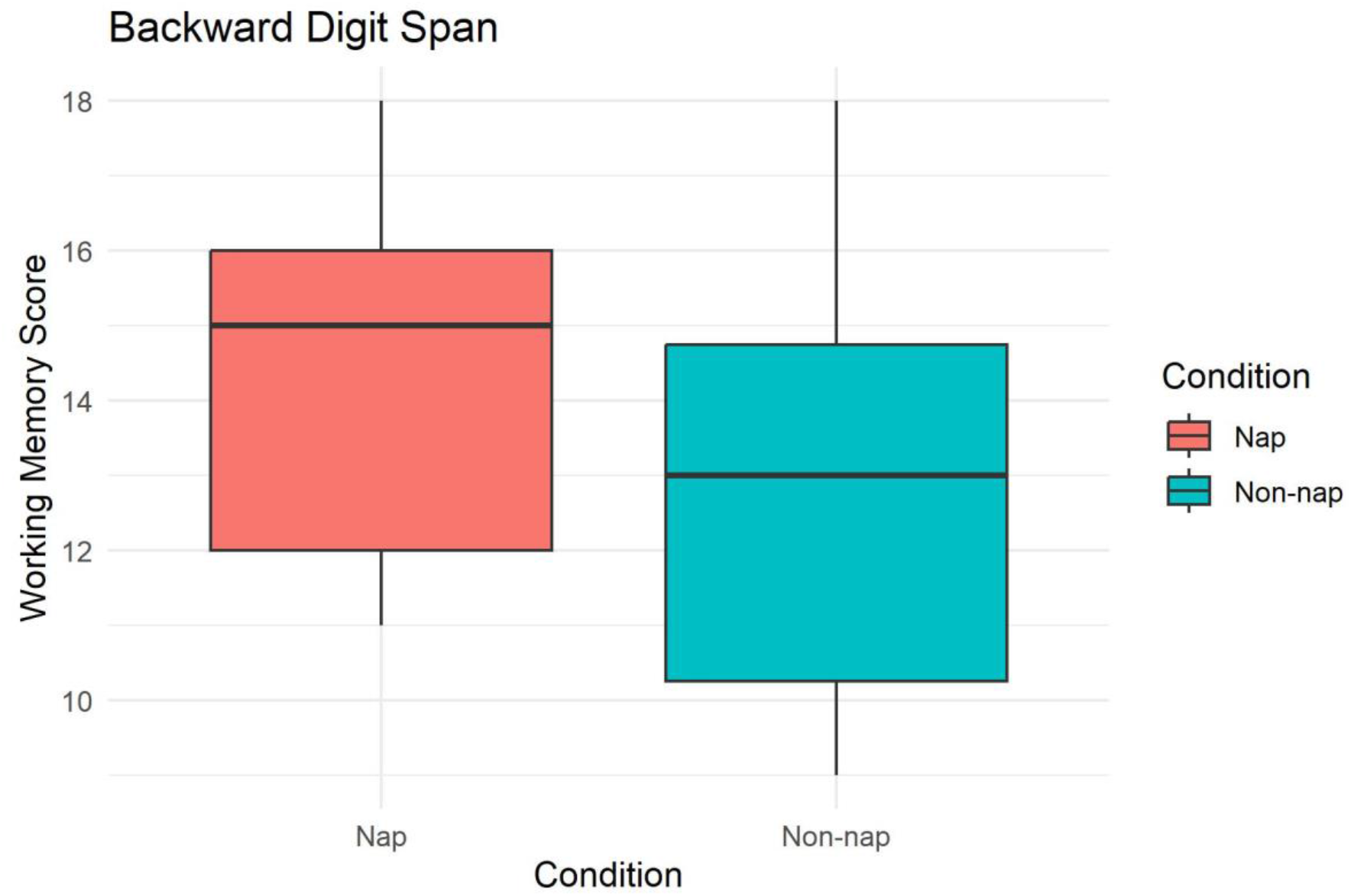
Working Memory Assessment using Backward Digit Span Task. The effect size (Hedges’ g = 0.459), measured as Hedges’ g, indicated a medium-sized effect, underscoring the meaningfulness of the observed improvement. The data highlight the potential cognitive benefits of a daytime nap on working memory abilities.

The significant improvement in Backward Digit Span performance with intervention suggests a positive effect on working memory. The observed effect size (Hedges’ g = 0.459) indicates a medium-sized effect, emphasizing the practical significance of the intervention in enhancing working memory function.

### 3.3. Attention

Based on the non-normality of the data, we performed Wilcoxon signed-rank tests to compare Stroop test scores. The results revealed significant differences in Paper-Pencil Stroop test scores between the two conditions for all three trials: the Control Trial (p = 0.006), Congruent trials (p = 0.009), and Incongruent trials (p = 0.005) (Table 3) (Figure 2). Despite the absence of a significant distinction in participants’ accuracy between the two conditions (Table 3), these results indicate that taking a nap had a noteworthy influence on Stroop test performance (Reaction Time), resulting in noticeable alterations in cognitive processes associated with attention and interference during the execution of the Stroop task.

**Table 3.** Results of the Stroop task Reaction Time.

| Trials |  | Mean | SD | p-value |
| --- | --- | --- | --- | --- |
| Control | Nap | 106.10 | 8.65 | 0.006 |
|  | Non-nap | 111.60 | 7.05 |  |
| Congruent | Nap | 87.50 | 4.88 | 0.009 |
|  | Non-nap | 91.50 | 3.77 |  |
| Incongruent | Nap | 171.90 | 8.31 | 0.005 |
|  | Non-nap | 182 | 7.28 |  |
| Error | Nap | 0.7 | 0.675 | 0.821 |
|  | Non-nap | 0.8 | 0.789 |  |

**Figure 2.**
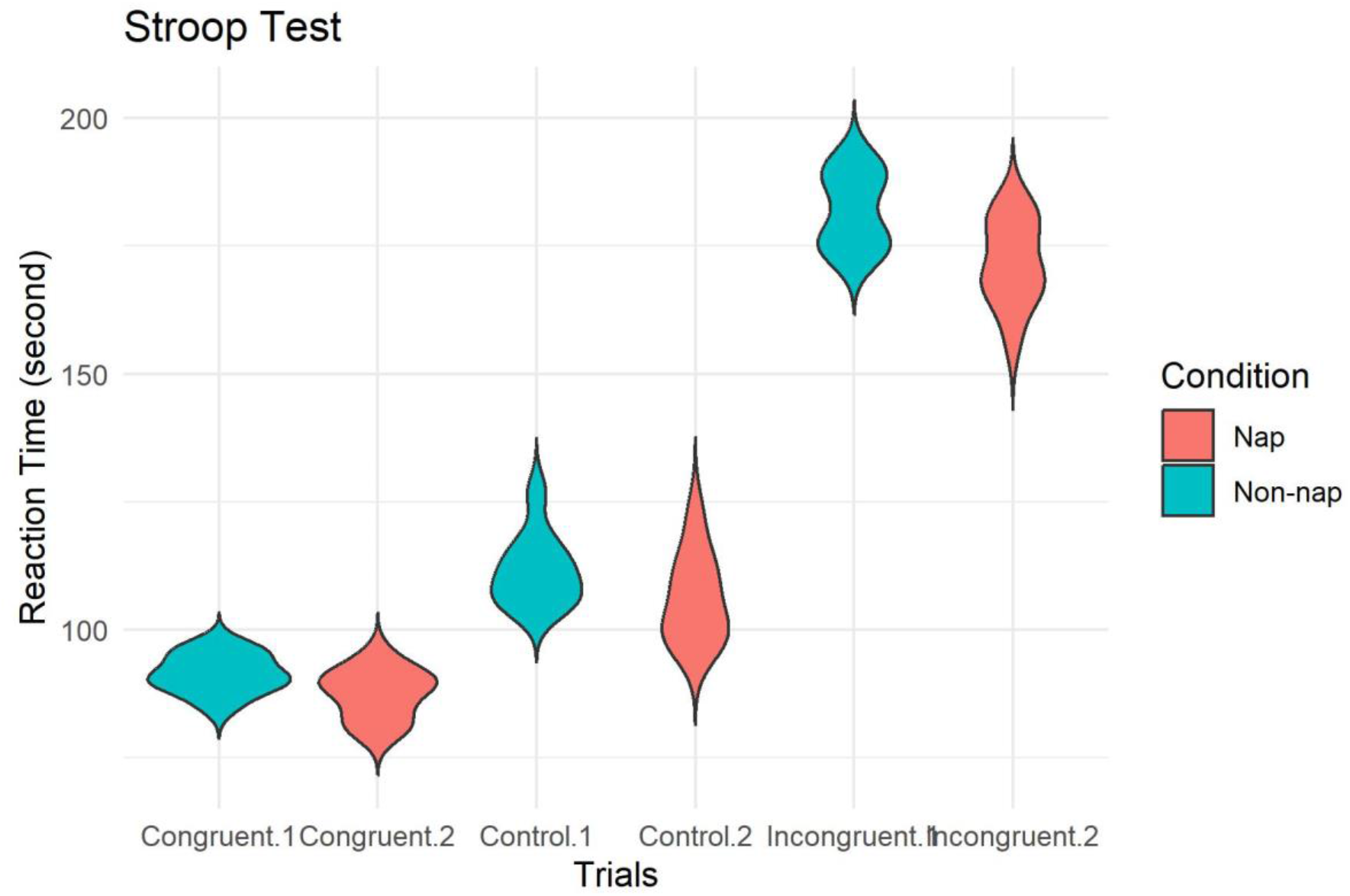
Attention Assessment using Stroop Test. In all three Stroop task trials, participants exhibited significantly improved reaction time in the nap condition compared to the no-nap condition (p < 0.05).

### 3.4. Recognition Memory

The statistical analysis using the Wilcoxon signed-rank tests revealed statistically significant changes in one subscale between the two experimental conditions. A significant difference was observed in the False Alarm scores, yielding a p-value of 0.015 at a significance level of α = 0.05. Nonetheless, there were no significant differences noted in the Hit scores (p = 0.173) (Table 5) (Figure 3).

**Table 5.**
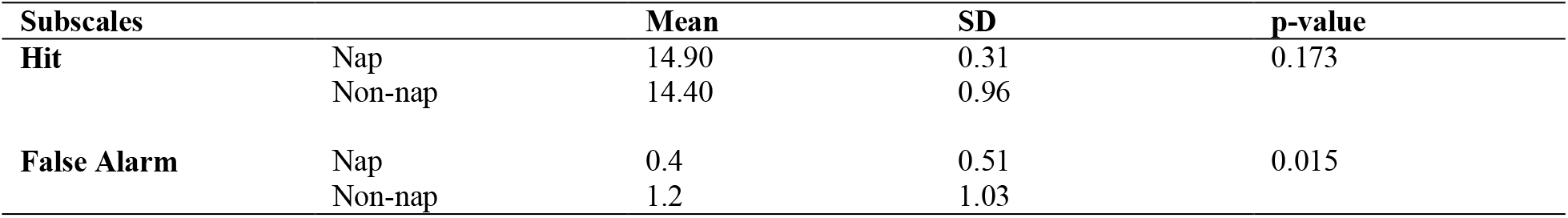
Results of Auditory Verbal Learning Test.

**Figure 3.**
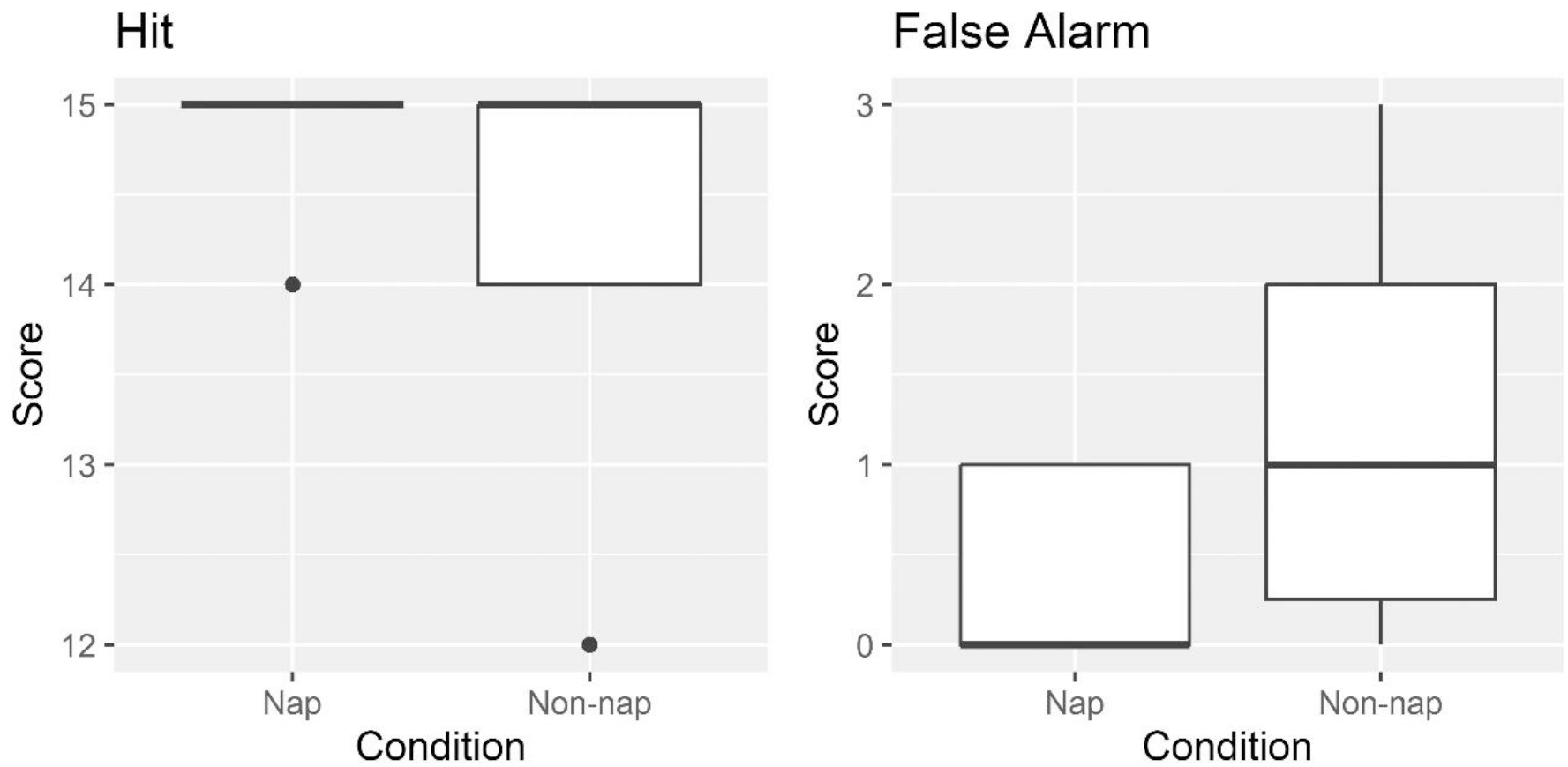
Recognition Memory Assessment using AVLT. While the hit scores did not exhibit a significant difference between the two conditions, participants’ performance on the nap condition demonstrated a statistically significant divergence in false alarm performance compared to the no-nap condition (p < 0.05).

## 4. Discussion

The primary aim of this study was to investigate the impact of daytime napping on attention, working memory, and recognition memory. Our findings lend support to the hypothesis that a daytime nap can significantly improve these cognitive functions. Cognitive functions were compared between the nap and non-nap conditions. Each participant was subjected to cognitive assessments in both conditions in order to minimize the between-group variation.

We found a significant difference in working memory performance between the two conditions. Participants performed significantly better when they napped in comparison with the non-nap condition. Notably, the effect size observed in our study was of medium size, emphasizing the practical significance of the findings. This finding is consistent with previous research suggesting that sleep, even for brief periods such as naps, can improve working memory performance (11, 35).

Naps taken by young adults, especially if they are of considerable duration, encompass both Non-Rapid Eye Movement (NREM) and Rapid Eye Movement (REM) episodes (17). On the other hand, naps among older adults are characterized by a prevalence of lighter NREM stages, a brief period of slow wave sleep (SWS), and, less frequently, REM sleep (18). Previous research on naps and nocturnal sleep revealed an elevation in mediated parasympathetic activity during the transition from wakefulness to NREM sleep (36, 37). These findings indicate a shift in the autonomic nervous system (ANS) from a state of sympathetic regulation to parasympathetic control as one moves from being awake to entering sleep.

As sleep ensues, there is a noticeable prevalence of parasympathetic dominance in comparison to the waking state, leading to the characterization of nighttime sleep as a “cardiovascular holiday” (38, 39). The overall balance between the parasympathetic and sympathetic branches of the autonomic nervous system holds significance for both health and cognitive function.

In line with current findings, Chen et al. (2020) observed that heightened parasympathetic activity during daytime naps could potentially support the enhancement of working memory (WM) (40).

Given that the primary motivation for daytime napping among healthy young adults appears to be restoration (41), previous results suggest that daytime naps might offer a compensatory protective period for the cardiovascular system (42). Daytime naps could be viewed as brief interludes that provide a “mini cardiovascular respite,” which holds promise for positively impacting cognitive functions.

Additionally, we delved into the influence of daytime naps versus wakefulness on attention performance, gauged through a conventional paper-pencil test. Our findings unveil a substantial contrast between the two conditions, indicating that the participant’s state (nap vs. wakefulness) likely exerts an impact on attention processing effectiveness.

Sleep, particularly daytime napping, has been firmly established as a pivotal element in maintaining alertness (43). Conversely, the negative impact of even short-term sleep deprivation on attention is well documented. Extended periods of wakefulness can lead to a decline in attention and other cognitive functions (5, 44). Previous research has demonstrated that napping during night shifts improves cognitive performance in a delayed manner, especially attention (45). Furthermore, fatigue and decreased alertness from prolonged wakefulness could also explain the weaker attention performance in the wake condition of our study. In line with our outcomes, Takahashi et al. demonstrated that a post-lunch nap at 12:30 h substantially improved alertness in individuals who had enjoyed 7 hours of sleep (46).

Furthermore, we probed the potential correlation between daytime napping and recognition memory. Although most prior studies involved participants learning prior to a nap and subsequently undergoing recognition memory assessments post-nap, our study focused on the recognition memory of learned words following a nap. Our investigation revealed that participants in a nap state exhibited superior performance in the recognition memory task. While no significant divergence was noted regarding the Hit subscale between the two conditions, a notable difference emerged in False alarms. Consistent with this outcome, previous research conducted in laboratory settings has shown that napping after learning aids in the consolidation of memory, while napping before learning facilitates the encoding of new information (19, 20, 21, 22). In harmony with our findings, certain previous studies have illuminated the favorable influence of daytime napping on recognition memory (47, 48, 49, 50, 51).

Despite the promising outcomes, it is essential to acknowledge the limitations of our study. One notable limitation is the relatively modest sample size, which may influence the generalizability of our findings. While the effect size suggests a meaningful improvement in working memory after a nap, future research with larger and more diverse participant cohorts is warranted to confirm and extend our results.

Another limitation pertains to our methodology for tracking naps. We did not utilize tools such as actigraph or polysomnography (PSG) to precisely monitor participants’ napping behavior. This could introduce some variability in nap duration, depth, and structure. While our study design aimed to replicate real-world conditions of daytime napping, incorporating more comprehensive tracking methods into future research could offer a more detailed understanding of the relationship between nap characteristics and cognitive outcomes.

In conclusion, our study provides valuable insights into the potential cognitive advantages of daytime napping, emphasizing the importance of sleep in promoting optimal cognitive functioning. As we move forward, a comprehensive understanding of the nuances surrounding daytime napping will undoubtedly contribute to the development of targeted interventions for various settings, from educational environments to workplaces, ultimately benefiting overall cognitive health and performance.

## Data Availability

The data and codes used in this study are available upon request. Interested parties may contact
Shahab Zare, at for access to the data and code

## 5. Declarations

### Author Contributions

Shahab Zare contributed to conceptualization, data collection, data analysis, code development, and manuscript writing. Javad Setareh provided crucial guidance and oversight throughout the project, lending expertise in sleep medicine, and facilitated the experimental environment in a sleep department of Zare Hospital. Fariborz Dortaj’s contributions included insightful discussions that contributed to project development and data analysis. All authors reviewed and approved the final version of the manuscript.

### Data and Code Availability

The data and code used in this are available upon request. Interested parties may contact Shahab Zare, at for access to the data and code.

### Clinical Trial Registration

The present study was prospectively registered with the Iranian Registry of Clinical Trials (IRCT) under registration number 66862. Date of registration: 12/31/2022

### Consent to participate

Participants were fully informed of the study’s purpose, procedures, risks, and benefits before participation. Informed consent was obtained before data collection, with emphasis on voluntary participation and the right to withdraw without consequences.

